# Low-Density Lipoprotein Cholesterol and Dementia Risk: Integrating Mendelian Randomization and Target Trial Emulation Within the Heart–Brain Axis

**DOI:** 10.64898/2026.06.10.26355413

**Authors:** Kudakwashe Mukumbi, Yanrong Liu, Zixin Shi, Edward Liu, Aili Toyli, Guang-Uei Hung, Qing-Hui Chen, Qiuying Sha, Pai-Yi Chiu, Weihua Zhou

## Abstract

**Background:** The heart–brain axis links cardiovascular and neurodegenerative disease through shared vascular and inflammatory mechanisms. Although low-density lipoprotein cholesterol (LDL-C) is an established causal factor in atherosclerotic cardiovascular disease (ASCVD), its relationship with dementia remains uncertain, with midlife elevations associated with increased risk but late-life associations often appearing null or inverse. To address this cholesterol paradox, we integrated mendelian randomization (MR) with an active-comparator new-user target trial emulation.

**Methods:** We applied a triangulated causal inference framework integrating two-sample MR with observational target trial emulation. Genetic variants associated with LDL-C were used as instrumental variables to evaluate Alzheimer’s disease (AD), Dementia with Lewy bodies (DLB), Frontotemporal dementia (FTD), and any dementia (AnyDem), with causal estimates derived using inverse-variance weighted models and sensitivity analyses for heterogeneity and pleiotropy. In parallel, an active-comparator new-user design compared statin versus ezetimibe initiation among adults aged ≥60 years using propensity score (PS) overlap weighting and Cox proportional hazards models to evaluate cardiovascular and dementia outcomes.

**Results:** Genetically predicted LDL-C was associated with increased risk of DLB (OR 1.65, 95% CI 1.30–2.10; p<0.001), but not AD or AnyDem; FTD estimates were inconsistent. Sensitivity analyses suggested heterogeneity and possible pleiotropy for DLB. In the observational analysis (n=6,977), statin initiation was associated with higher risks of ASCVD (HR 1.26, 95% CI 1.11–1.45) and AnyDem (HR 1.66, 95% CI 1.16–2.38), although estimates attenuated after lipid adjustment and lagged analyses, suggesting residual confounding, treatment selection, and reverse causation in late-life observational associations.

**Conclusions:** These findings suggest that LDL-C reflects accumulated vascular and metabolic risk rather than a direct causal driver of AD or overall dementia, although a subtype-specific association was observed for DLB. Late-life associations appeared influenced by timing, reverse causation, and treatment selection, warranting cautious interpretation.

## Introduction

CVD remains the leading cause of mortality worldwide and an increasingly important contributor to age-related morbidity^1^. Its overlap with neurodegenerative disorders, particularly dementia, has prompted growing interest in the heart–brain axis, in which vascular injury, inflammation, metabolic dysfunction, and shared cardiometabolic risk factors may connect cardiovascular and neurodegenerative processes^2-5^. Within this framework, LDL-C is a particularly important but unresolved factor. Although LDL-C has an established causal role in ASCVD^6^, its relationship with dementia remains unclear. Epidemiologic studies have reported inconsistent associations between LDL-C and dementia^7, 8^, and MR studies have not consistently supported a direct causal effect of LDL-C on neurodegeneration^9^. This contrast raises a central question: whether LDL-C contributes to dementia through shared vascular pathways, or whether observed associations mainly reflect timing, disease progression, and bias.

A major challenge in interpreting LDL-C and dementia associations is that cholesterol may carry different meanings across the life course. Elevated cholesterol in midlife has been associated with increased dementia risk^10^, whereas late-life cholesterol levels are often reported as null or inversely associated with dementia^8, 11^. Longitudinal evidence further suggests that cholesterol decline may precede dementia onset^12^, indicating that late-life lipid levels may be influenced by subclinical disease, frailty, metabolic changes, or reverse causation rather than representing a direct causal exposure. This pattern, often described as the cholesterol paradox, is further complicated by evidence of non-linear associations, in which both elevated and very low cholesterol levels may be linked to adverse cognitive outcomes in older or preclinical populations^13-15^. Consistent with this interpretation, systematic reviews suggest that midlife cholesterol is a more reliable predictor of dementia risk than late-life cholesterol levels^16^.

Clarifying this distinction is clinically important because LDL-C is modifiable and lipid-lowering therapies are widely used in older adults, yet their relevance to dementia prevention remains uncertain^17-19^. Observational treatment comparisons in late life may be particularly vulnerable to confounding by indication, treatment selection, and reverse causation, while genetic analyses capture lifelong lipid-related liability rather than short-term therapeutic effects. To address these complementary questions, we integrated two-sample Mendelian randomization with an active-comparator new-user target trial emulation. The MR analysis evaluated whether lifelong genetically predicted LDL-C is associated with dementia outcomes, while the target trial emulation compared late-life initiation of statin versus ezetimibe therapy in relation to cardiovascular and dementia outcomes. By combining these approaches, this study aimed to distinguish potential biological effects of LDL-C from late-life treatment-related and observational processes within the heart–brain axis.

## Materials and Methods

### Study Design

We conducted a triangulated causal inference study to evaluate the relationship between LDL-C, cardiovascular disease, and dementia within the heart–brain axis. To guide the study design, we developed a conceptual and analytical framework that distinguishes the established causal pathway linking LDL-C to ASCVD from more complex and potentially indirect pathways linking LDL-C to dementia. This framework also illustrates how Mendelian randomization and target trial emulation provide complementary evidence on lifelong lipid-related liability and late-life treatment associations, respectively (Figure 1).

**Figure 1.**
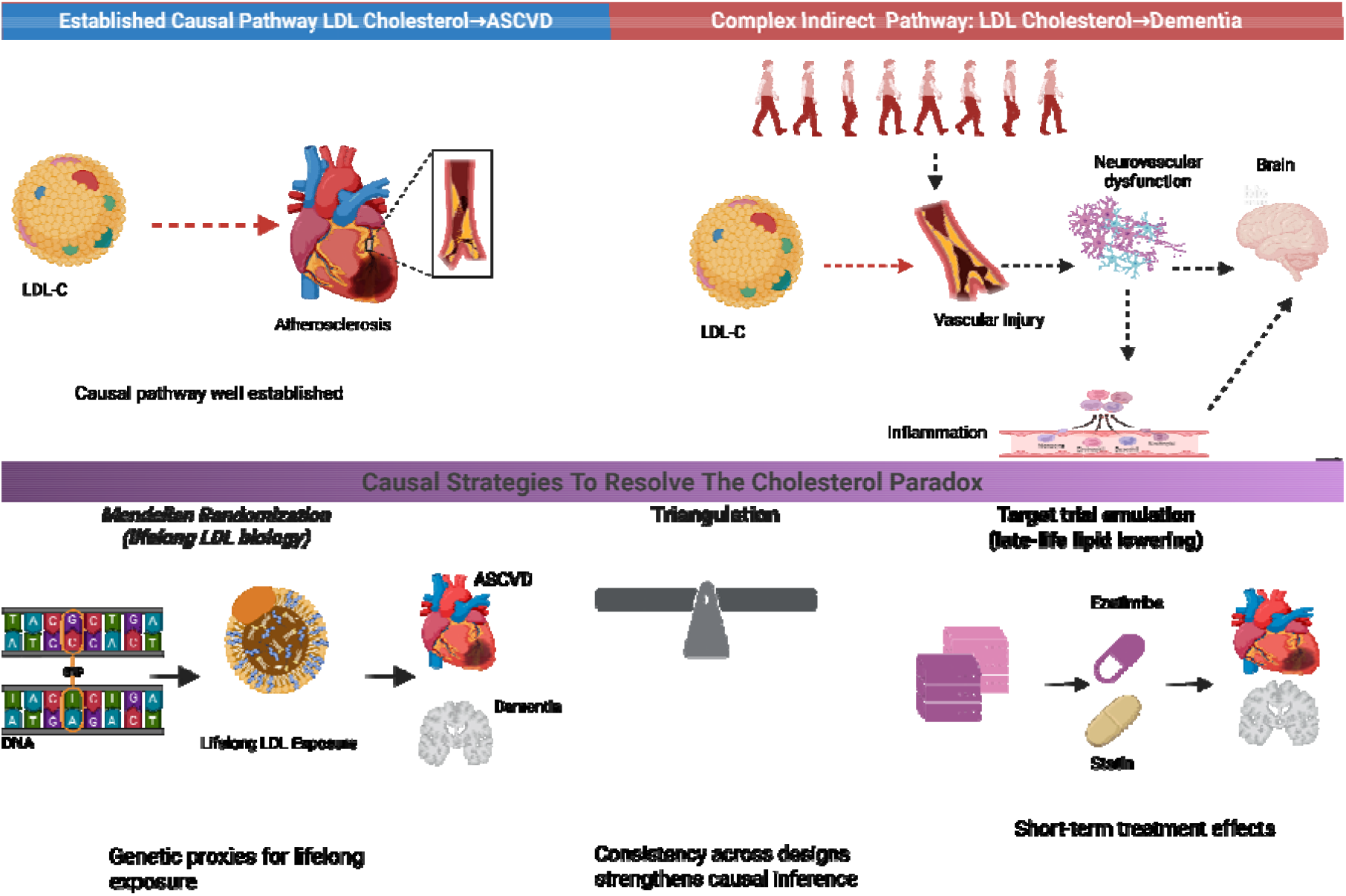
Conceptual heart-brain axis framework underlying the cholesterol paradox. Solid arrows represent hypothesized causal pathways linking lifelong LDL-C exposure to vascular injury and dementia. Dashed arrows represent non-causal pathways, Including confounding and reverse causation driven by subclinical disease and frailty. Red dashed arrows indicate reverse causation pathways influenced by disease progression, whereas black dashed arrows denote indirect or observational associations. The framework distinguishes lifelong biological exposure (captured through Mendelian randomization) from fete-life treatment effects (evaluated using target trial emulation), highlighting how bias may influence observed associations

We then implemented two complementary analytical components. First, two-sample Mendelian randomization was used to estimate the association between genetically predicted LDL-C and dementia outcomes, including AD, DLB, FTD, and any dementia. In this analysis, genome-wide significant SNPs associated with LDL-C were selected as genetic instruments and applied to independent GWAS datasets for dementia outcomes. Second, the observational component emulated an active-comparator new-user target trial using the All of Us Research Program, focusing on adults aged ≥60 years who initiated statin or ezetimibe therapy. Individuals with prevalent dementia or prior ASCVD were excluded, and at least 365 days of continuous pre-index observation was required. Treatment assignment was defined at initiation, and participants were followed until outcome occurrence, censoring, or the end of available observation. Propensity score-based overlap weighting was used to improve covariate balance between treatment groups.

Together, these two approaches were designed to separate lifelong biological associations of LDL-C from late-life treatment-related associations that may be influenced by treatment selection, residual confounding, frailty, or reverse causation. The overall workflow, including cohort selection, exposure definition, and outcome evaluation across both analytical components, is summarized in Figure 2.

**Figure 2.**
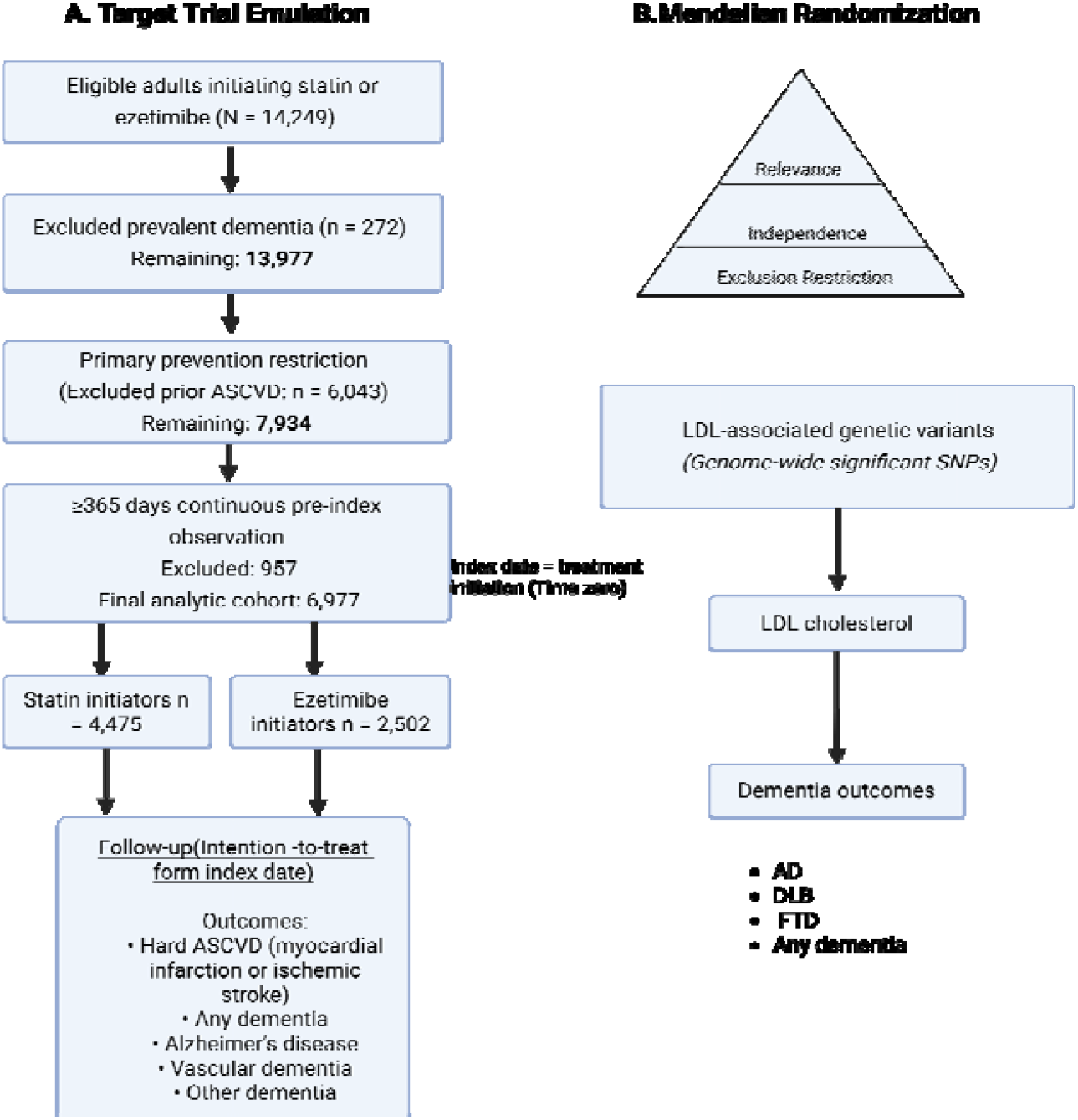
Study design integrating target trial emulation and Mendelian.

### Data Sources

MR analyses utilized GWAS summary statistics from the MRC-IEU OpenGWAS platform^20^. Genetic instruments for LDL-C were obtained from the Global Lipids Genetics Consortium^21^, and outcome data were obtained from multiple GWAS consortia and biobank studies, as detailed in Table 1. The datasets largely comprised populations of European ancestry, reducing potential bias due to population stratification.

**Table 1.**
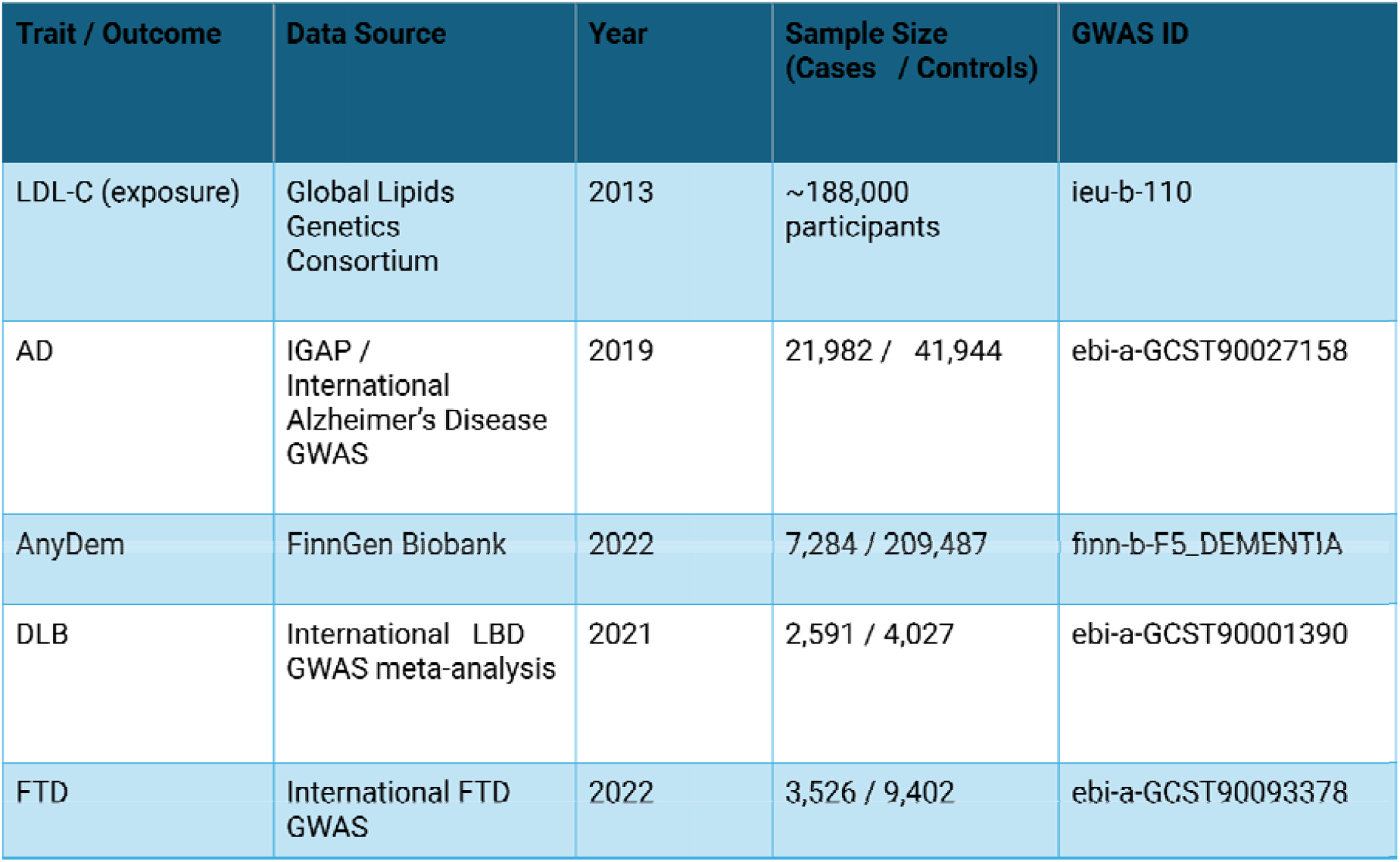
Summary of GWAS datasets used for cholesterol exposure and neurodegenerative outcomes.

| Trait / Outcome | Data Source | Year | Sample Size (Cases / Controls) | GWAS ID |
| --- | --- | --- | --- | --- |
| LDL-C (exposure) | Global Lipids Genetics Consortium | 2013 | ~188,000 participants | ieu-b-110 |
| AD | IGAP / International Alzheimer's Disease GWAS | 2019 | 21,982 / 41,944 | ebi-a-GCST90027158 |
| AnyDem | FinnGen Biobank | 2022 | 7,284 / 209,487 | finn-b-F5_DEMENTIA |
| DLB | International LBD GWAS meta-analysis | 2021 | 2,591 / 4,027 | ebi-a-GCST90001390 |
| FTD | International FTD GWAS | 2022 | 3,526 / 9,402 | ebi-a-GCST90093378 |

### Instrumental Variable Selection

SNPs associated with LDL-C at genome-wide significance (p < 5 × 10_DD_) were selected as instrumental variables^22, 23^. Linkage disequilibrium clumping was applied to ensure independence between SNPs. Where required, proxy SNPs in high linkage disequilibrium (r^2^_≥_ 0.8) were used to maximize overlap between exposure and outcome datasets. After harmonization and removal of palindromic SNPs, final instrument counts were 160 for AD, 150 for DLB, 137 for FTD, and 165 for AnyDem.

### Statistical Analysis

Causal effects were estimated using the inverse-variance weighted (IVW) method^22^. Sensitivity analyses included MR-Egger regression, weighted median, and weighted mode approaches^22-24^. Heterogeneity was assessed using Cochran’s Q statistic, and directional pleiotropy was evaluated using the MR-Egger intercept. Additional sensitivity for DLB included leave-one-out testing, single-SNP analyses, and MR-PRESSO to assess the influence of individual variants and potential outlier effects^23^. Effect estimates were reported as ORs with 95% CI.

Observational Analysis (Target Trial Emulation)

### Data Source and Eligibility Criteria

The observational analysis emulated a target trial^25^ using data from the All of Us Research Program^26^. Clinical data were structured using the Observational Medical Outcomes Partnership (OMOP) Common Data Model^26^. Outcomes and baseline comorbidities were defined using standardized OMOP concept sets derived from Systematized Nomenclature of Medicine -- Clinical Terms (SNOMED) and International Classification of Diseases (ICD) mappings through the concept_ancestor table to ensure consistent phenotype definitions. Eligible participants were adults aged ≥60 years initiating statin or ezetimibe therapy with at least 365 days of continuous observation prior to treatment initiation. Participants were classified based on their first eligible prescription, and individuals with prevalent dementia or prior ASCVD were excluded from incident outcome analyses.

### Baseline Covariates

Baseline covariates assessed in the year prior to treatment initiation included demographic characteristics, healthcare utilization, smoking status, comorbidities, and concomitant medications. Baseline lipid measures were included in sensitivity analyses.

### Propensity Score Modeling and Overlap Weighting

PSs were estimated using logistic regression including demographic variables (age, sex, race, and ethnicity), baseline comorbidities (type 2 diabetes, hypertension, hyperlipidemia, chronic kidney disease, heart failure, atrial fibrillation, obesity, and chronic obstructive pulmonary disease), healthcare utilization, and smoking status measured during the 365-day pre-index period. Education level was not consistently available in the dataset and was therefore not included. Overlap weighting was applied to minimize confounding, assigning weights of (1 – PS) for statin users and PS for ezetimibe users, thereby targeting a population in clinical equipoise. Covariate balance after weighting was assessed using standardized mean differences (SMDs), with values <0.1 indicating adequate balance between treatment groups.

### Statistical Analysis and Outcomes

Treatment assignment was defined using OMOP drug exposure data, with the index date corresponding to the first eligible prescription after age 60. Follow-up continued until outcome occurrence, censoring, or end of observation. The primary endpoint was hard ASCVD, and dementia outcomes were defined using standardized OMOP concept definitions.

Follow-up began at the index date and continued until the first occurrence of the outcome, death, loss to follow-up, or end of the study period. Overlap-weighted Cox proportional hazards models were used to estimate hazard ratios (HRs) and 95% CI. Sensitivity analyses included lipid-adjusted models incorporating baseline LDL-C (with additional adjustment for HDL-C and triglycerides where available) and lagged analyses (1-year and 2-year lags) to assess robustness and reduce potential reverse causation. A negative control outcome was used to evaluate residual confounding.

## Results

Genetically predicted LDL-C was associated with increased risk of DLB (IVW OR 1.652, 95% CI 1.298–2.103; p<0.001), with no evidence of a causal association for AD (OR 0.965, 95% CI 0.890–1.047) or AnyDem (OR 1.086, 95% CI 0.912–1.292); FTD estimates were inconsistent across methods (Table 2, Figure 3).

**Table 2.** MR estimates of LDL-C on dementia outcomes.

| Outcome | Method | SNPs | OR | 95% CI | p-value |
| --- | --- | --- | --- | --- | --- |
| AD | IVW | 160 | 0.965 | 0.890–1.047 | p = 0.389 |
|  | MR-Egger | 160 | 0.901 | 0.794–1.024 | p = 0.112 |
|  | Weighted median | 160 | 0.915 | 0.841–0.996 | p = 0.040 |
|  | Weighted mode | 160 | 0.909 | 0.846–0.976 | p = 0.010 |
| DLB | IVW | 150 | 1.652 | 1.298–2.103 | p < 0.001 |
|  | MR-Egger | 150 | 2.218 | 1.624–3.029 | p < 0.001 |
|  | Weighted median | 150 | 2.027 | 1.526–2.693 | p < 0.001 |
|  | Weighted mode | 150 | 1.664 | 1.280–2.164 | p < 0.001 |
| FTD | IVW | 137 | 1.708 | 1.057–2.761 | p = 0.029 |
|  | **MR-Egger** | **137** | **1.904** | **0.900–**<br>**4.030** | **p =**<br>**0.095** |
|  | **Weighted median** | **137** | **2.037** | **0.868–**<br>**4.784** | **p =**<br>**0.102** |
|  | **Weighted mode** | **137** | **1.823** | **0.783–**<br>**4.248** | **p =**<br>**0.166** |
| **AnyDem** | **IVW** | **165** | **1.086** | **0.912–**<br>**1.292** | **p =**<br>**0.356** |
|  | **MR-Egger** | **165** | **1.202** | **0.923–**<br>**1.566** | **p =**<br>**0.173** |
|  | **Weighted median** | **165** | **0.923** | **0.766–**<br>**1.112** | **p =**<br>**0.399** |
|  | **Weighted mode** | **165** | **0.930** | **0.783–**<br>**1.104** | **p =**<br>**0.405** |

**Figure 3.**
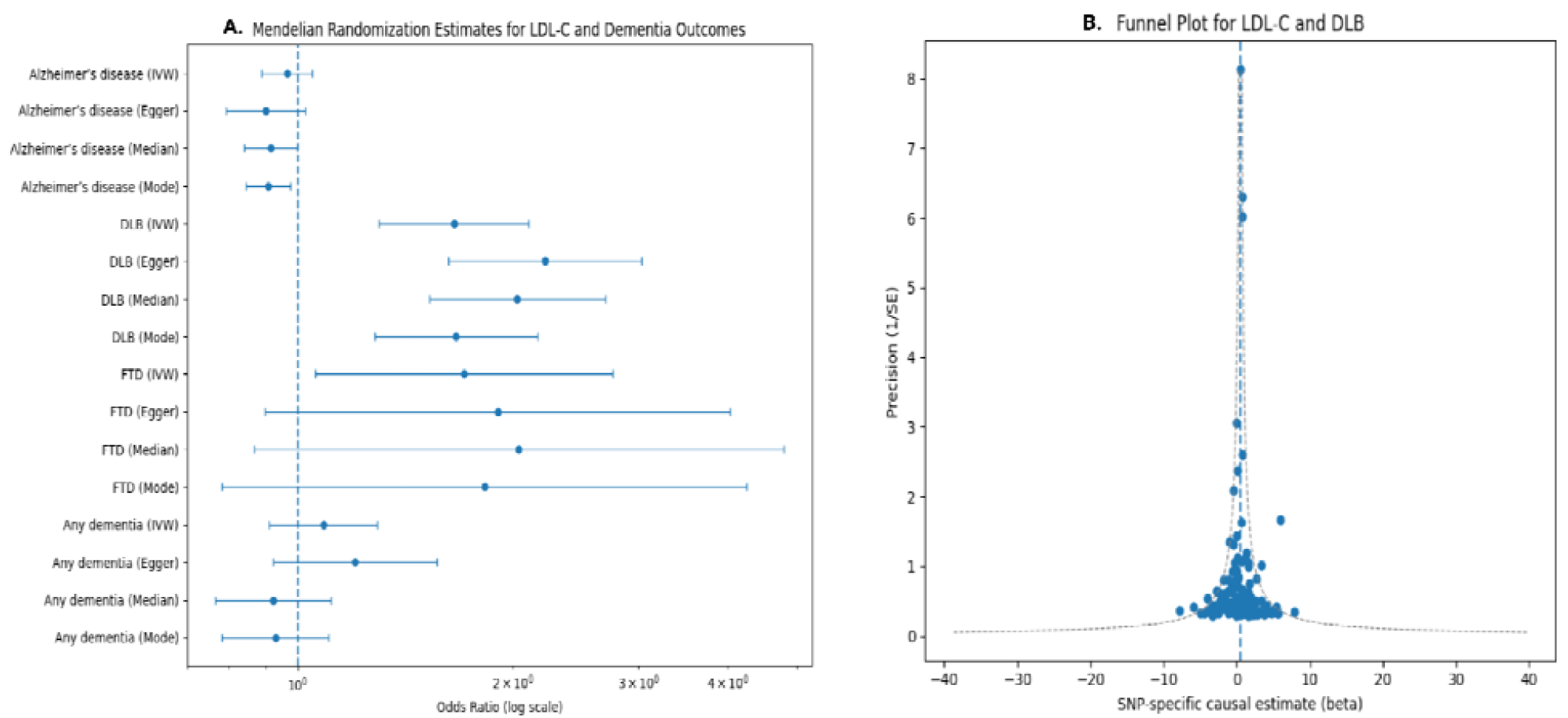
Mendellan Randomization Estimates for LDL cholesterol and Dementia outcomes. Forest plot showing the association between genetically predicted LDL-C and dementia outcomes across MR methods (A). Funnel plot assessing potential directional plelotropy for LDL-C and DLB (B)

Sensitivity analyses showed heterogeneity across SNP-specific estimates and suggested possible pleiotropy for DLB, with no evidence of directional pleiotropy for other outcomes (Figure 3).

In the observational analysis (n=6,977), substantial baseline differences were observed prior to weighting, with a maximum standardized mean difference of 0.940, indicating significant imbalance between treatment groups (Figure 4). After applying overlap weighting, covariate balance was achieved, with all SMDs <0.100 (Figure 4). The variables included in the balance diagnostics (Figure 4) correspond to all covariates used in the PS model, including demographic characteristics, comorbidities, healthcare utilization, and smoking status.

**Figure 4.**
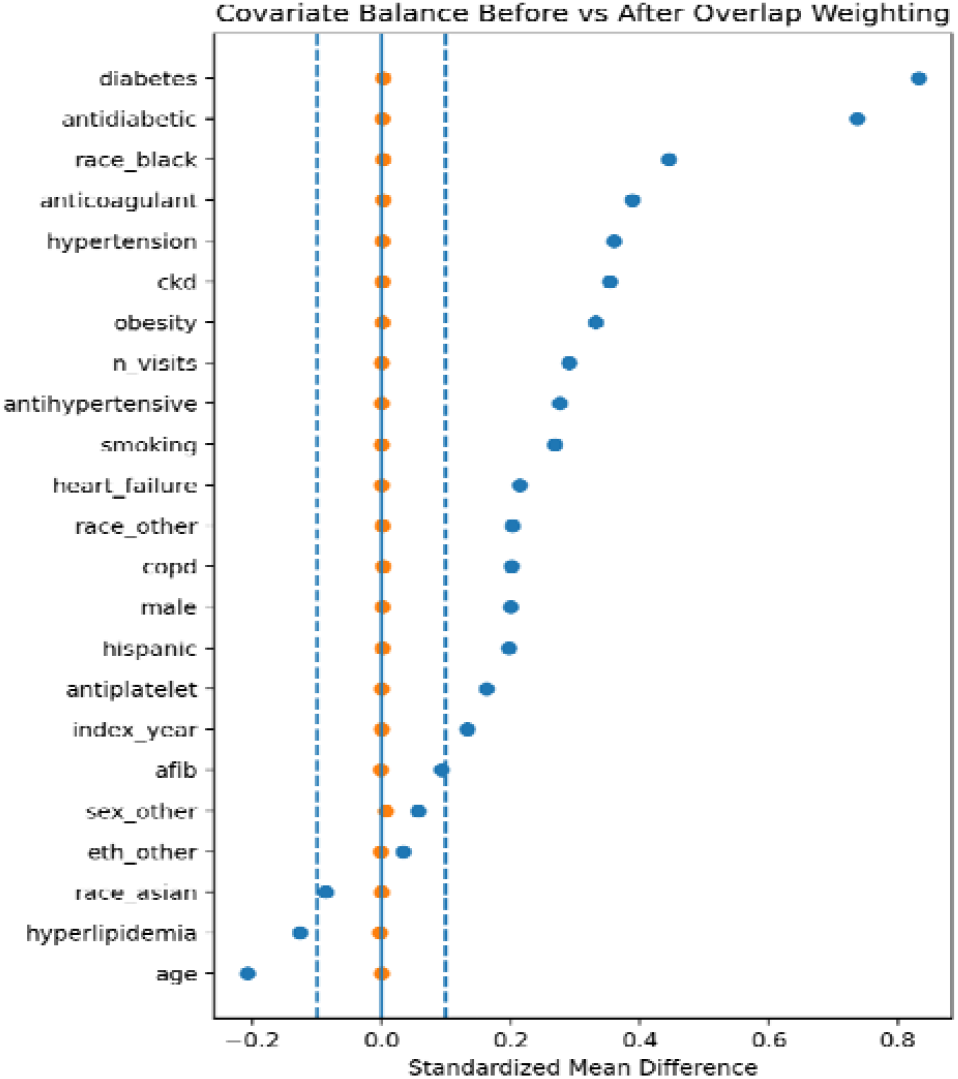
Covariate balance assessed using standardized mean differences before and after overlap weighting. Values below 0.1 Indicate adequate balance.

PS distributions showed limited overlap before weighting and improved overlap after applying overlap weighting (Figure 5).

**Figure 5.**
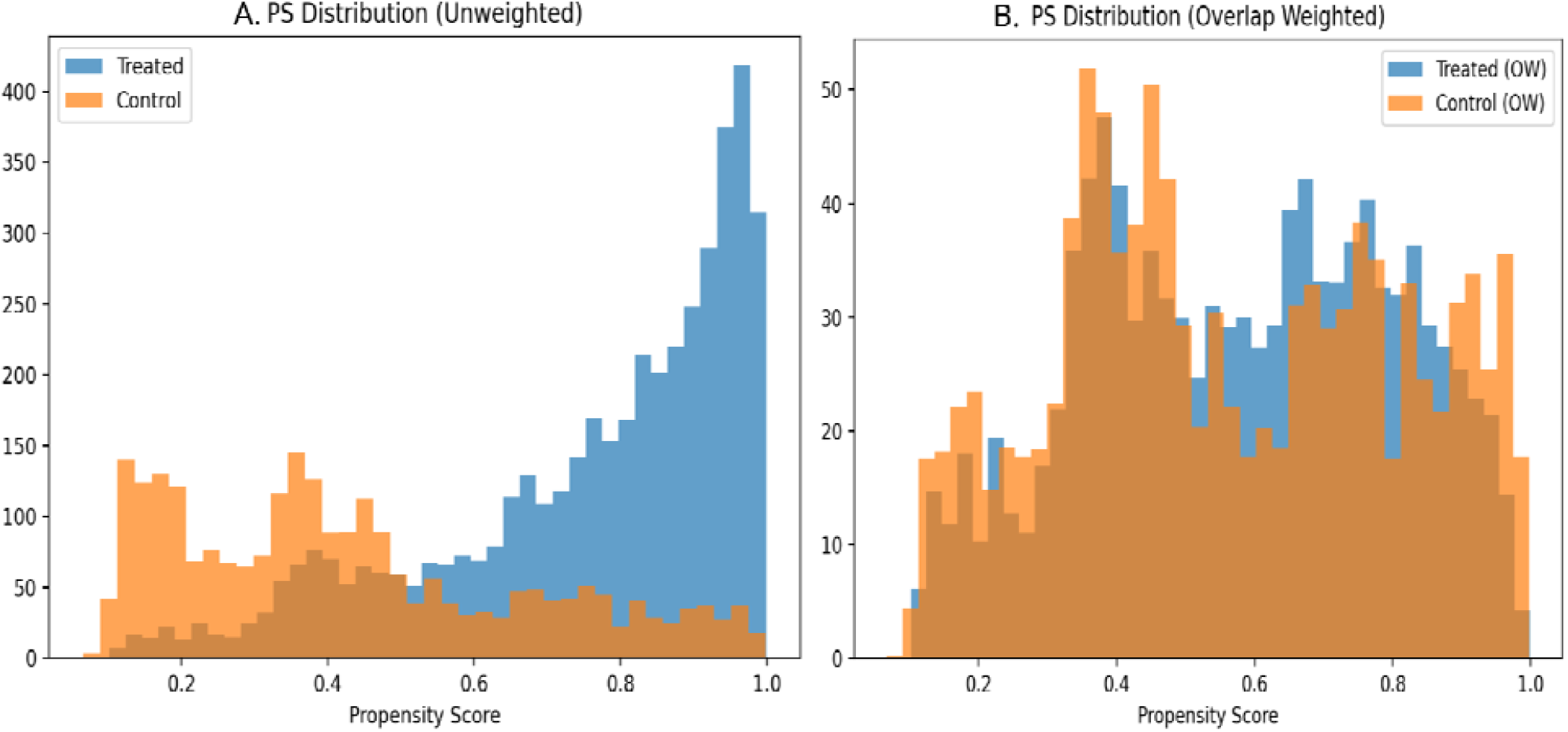
Propensity score distributions before and after overlap weighting. (A) Unweighted propensity score distributions for statin and ezetimibe initiators, showing limited overlap between treatment groups. (B) Overlap-weighted propensity score distributions demonstrating improved balance and overlap.

Sensitivity analyses showed that adjustment for baseline lipids did not materially change effect estimates compared with the primary analysis (Figure 6). Lagged analyses yielded less precise and partially attenuated estimates. At 1-year lag, several dementia outcomes had insufficient events or unstable estimates, limiting interpretability. At 2-year lag, effect estimates for both ASCVD (HR 1.193) and AnyDem (HR 1.458) moved closer to the null, with wider CIs indicating reduced precision.

**Figure 6.**
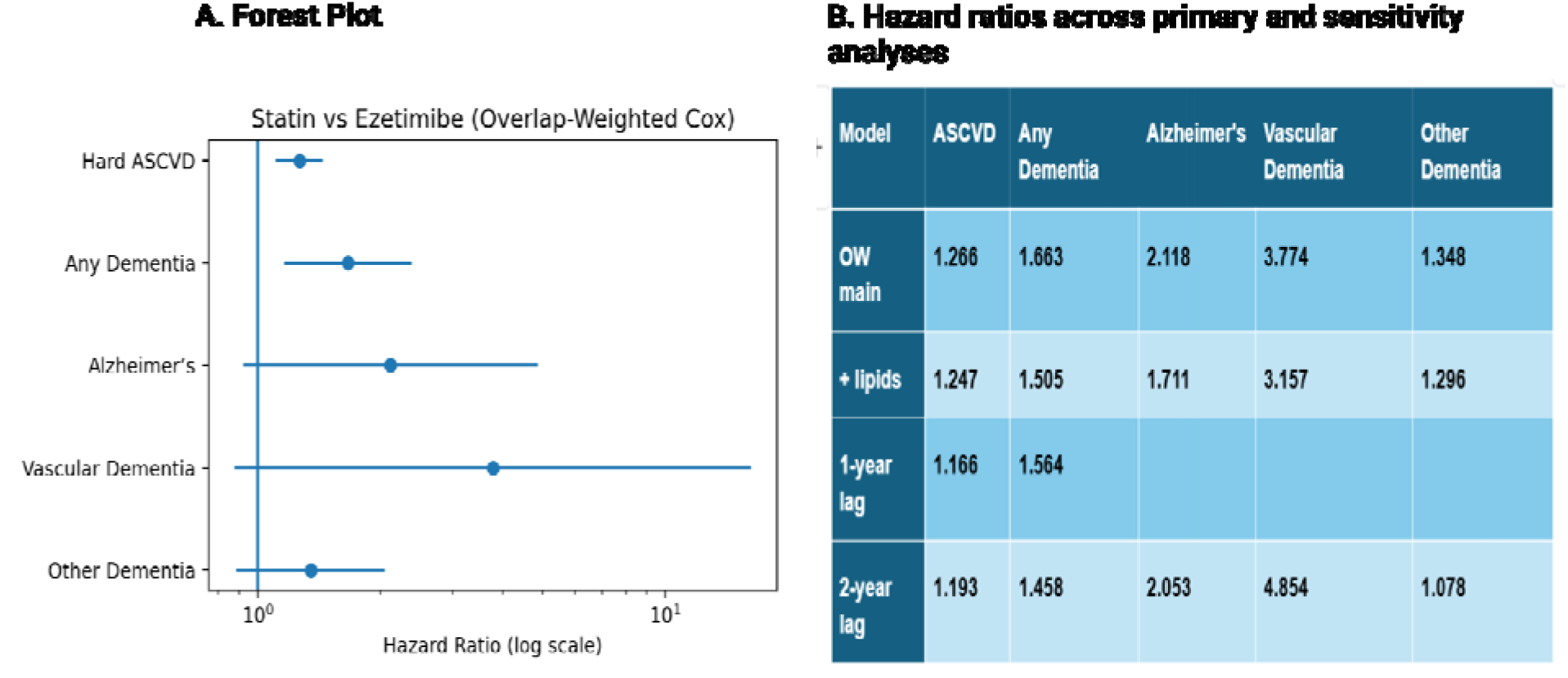
Primary and sensitivity analyses of cardiovascular and dementia outcomes. (A) Forest plot showing overlap-weighted hazard ratios and 95% confidence Intervals comparing statin versus ezetimlbe Initiation for ASCVD and dementia outcomes. Hazard ratios greater than 11ndicate higher risk associated with statin use relative to ezetimlbe. (B) Corresponding hazard ratios across primary, lipid-adjusted, and lagged analyses (1-year and 2-year). Blank cells indicate Insufficient event counts after applying lag restrictions.

### Discussion Principal Findings

In this triangulated causal inference study, genetically predicted LDL-C was associated with increased risk of DLB, suggesting a potential subtype-specific relationship between lipid metabolism and neurodegeneration. This finding is consistent with prior MR evidence linking higher LDL-C levels to DLB risk^27^. However, the DLB result should be interpreted cautiously because sensitivity analyses indicated heterogeneity and possible pleiotropy. In contrast, we did not observe consistent evidence supporting a causal association between LDL-C and AD or AnyDem, and the evidence for FTD was inconsistent across MR methods. These findings suggest that LDL-C is unlikely to act as a universal causal driver of dementia, although it may be relevant to specific dementia subtypes.

The MR findings provide context for interpreting the observational target trial emulation results. In overlap-weighted analyses, statin initiation was associated with higher risks of ASCVD and dementia compared with ezetimibe initiation, but these associations should not be interpreted as evidence that statins increase dementia risk. Although adjustment for baseline lipid measures did not materially change the estimates, lagged analyses produced less precise and partially attenuated associations. These patterns suggest that treatment selection, residual confounding, early outcome events, and reverse causation may have contributed to the observed late-life associations.

### Interpretation within the Heart–Brain Axis and Cholesterol Paradox

The contrast between lifelong genetic evidence and late-life treatment associations highlights the importance of exposure timing. Prior longitudinal studies have shown that cholesterol trajectories over time may be more informative for dementia risk than single late-life lipid measurements^10, 12^. In particular, declining cholesterol before dementia onset may reflect subclinical disease, frailty, metabolic changes, or reverse causation rather than a protective effect of higher late-life cholesterol. Evidence of non-linear associations, including inverted U-shaped relationships between cholesterol and cognitive outcomes, further suggests that both high and low cholesterol levels may have different meanings across stages of cognitive decline^14, 15^.

Within the heart–brain axis, LDL-C may contribute to neurodegeneration indirectly through vascular injury, inflammation, neurovascular dysfunction, and accumulated cardiometabolic burden^6, 17^. However, the present findings do not support a simple pathway in which higher LDL-C directly increases risk for all dementia outcomes. Instead, the results suggest a more nuanced interpretation: LDL-C is clearly relevant to cardiovascular disease, may have subtype-specific relevance to DLB, and may also serve as a marker of broader vascular and metabolic risk in late-life observational settings.

The target trial emulation results are consistent with this interpretation. Statin use in older adults has not been consistently associated with reduced dementia risk in prior studies^18, 28^ and pharmacoepidemiologic analyses of late-life medication use are particularly susceptible to reverse causation and time-related biases^29^. In this context, the higher dementia risk observed among statin initiators compared with ezetimibe initiators likely reflects differences in baseline risk, treatment selection, and disease timing rather than a sustained causal effect of statin therapy on dementia. These findings support continued use of lipid-lowering therapy for cardiovascular prevention, while suggesting that late-life initiation of lipid-lowering treatment should not be assumed to provide short-term dementia prevention benefits.

### Mechanistic Interpretation

The directed acyclic graph in Figure 7 provides a conceptual interpretation of these findings by distinguishing potential biological pathways from non-causal observational processes. LDL-related biology is positioned as an upstream contributor to vascular injury, which may indirectly influence neurodegenerative risk through vascular and inflammatory mechanisms. At the same time, late-life LDL-C measurements and treatment initiation may be influenced by subclinical disease, frailty, healthcare utilization, and treatment selection. These processes can create observational associations that do not necessarily reflect direct causal effects.

**Figure 7.**
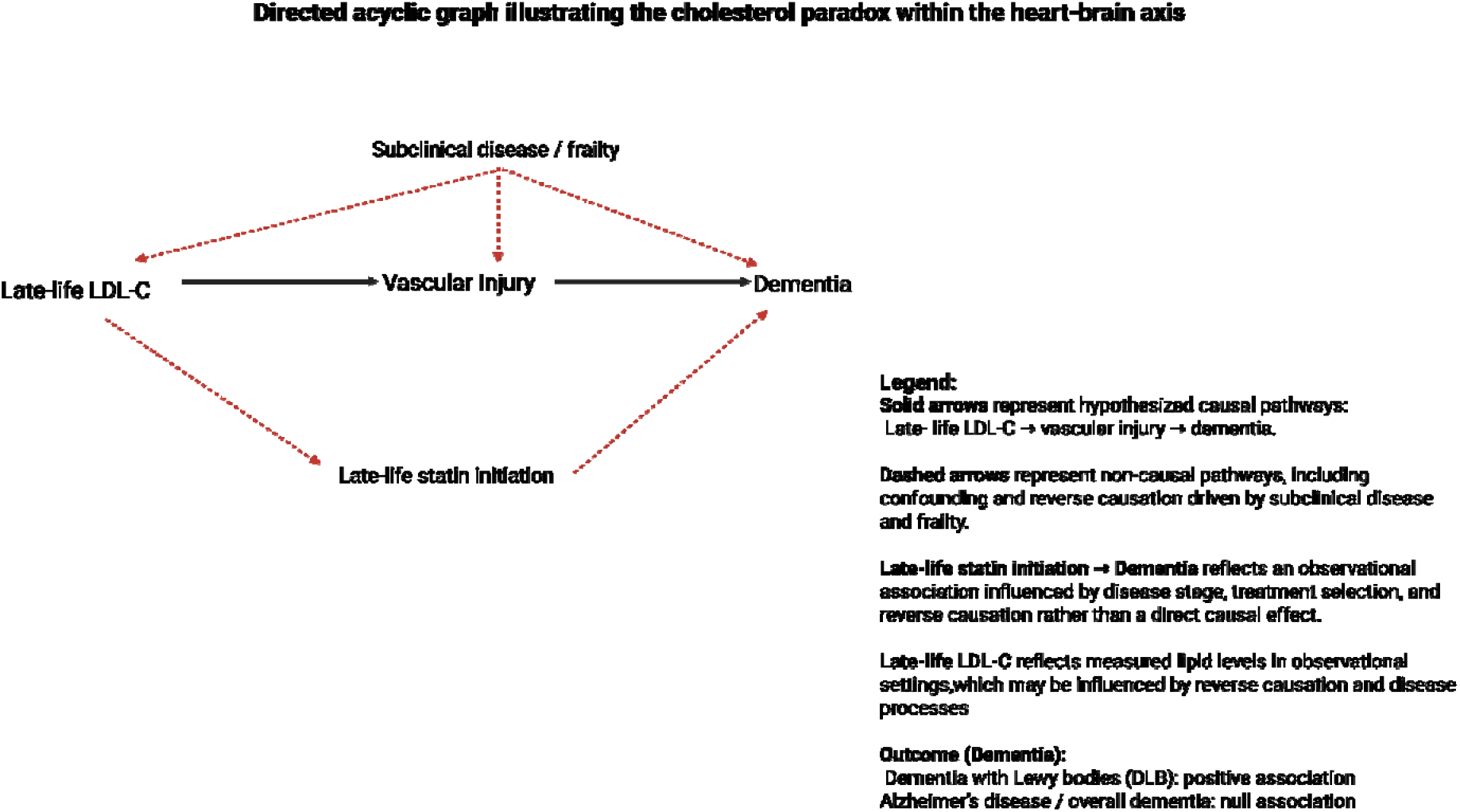
Conceptual Framework illustrating the cholesterol paradox within heart-brain axis.

This framework helps explain why midlife cholesterol may be associated with later dementia risk, whereas late-life cholesterol associations are often null, inverse, or inconsistent. It also helps reconcile the MR and target trial emulation findings: MR captures lifelong lipid-related genetic liability, whereas target trial emulation evaluates late-life treatment initiation in a real-world clinical setting. The combined evidence suggests that LDL-C is not a universal causal factor for dementia, but may contribute to dementia risk indirectly or in a subtype-specific manner, particularly for DLB.

## Limitations and Future Research

This study has several limitations. First, the DLB MR analysis showed evidence of heterogeneity and possible horizontal pleiotropy, which may affect causal interpretation despite sensitivity analyses. Second, MR estimates reflect lifelong genetic liability to LDL-C and may not directly correspond to the effects of lipid-lowering therapy initiated in late life^24, 30^. MR assumptions, including the absence of horizontal pleiotropy and valid exclusion restriction, cannot be fully verified. Third, although the target trial emulation used an active-comparator new-user design and overlap weighting, the observational analysis remains susceptible to residual confounding, reverse causation, and time-related biases^31, 32^. Fourth, dementia outcomes defined from EHR data may be affected by outcome misclassification, incomplete capture of diagnoses, and differences in healthcare utilization. Finally, the predominance of European ancestry populations in the GWAS datasets may limit generalizability to more diverse populations.

Future research should focus on subtype-specific mechanisms, particularly in Lewy body disease, and on clarifying non-linear and stage-dependent relationships between cholesterol and cognitive decline. Longitudinal studies with repeated lipid measurements, detailed medication histories, diverse populations, and well-characterized dementia subtypes will be important for distinguishing causal effects from disease-related changes and treatment-selection processes.

## Conclusion

Lifelong LDL-C exposure shows a subtype-specific association with dementia with Lewy bodies rather than a universal relationship with dementia. Late-life cholesterol levels and statin initiation were not associated with dementia risk, with attenuation after accounting for timing-related biases and reverse causation. These findings support a subtype-specific, indirect role of LDL-C and suggest that the cholesterol paradox reflects differences between lifelong biological exposure and late-life observational effects.

## Data Availability

The computer source code for this study is available at https://github.com/MIILab-MTU/HeartBrain_LDL_DementiaRisk. Individual-level data from the All of Us Research Program are available to approved researchers through the All of Us Researcher Workbench and cannot be shared directly by the authors. GWAS summary statistics used for the Mendelian randomization analyses were obtained from publicly available sources described in the manuscript.

## Non-standard Abbreviations and Acronyms

**Abbreviation Full Term**

AD: Alzheimer’s disease
AoU: All of Us Research Program
ASCVD: Atherosclerotic cardiovascular disease
BMI: Body mass index
CI: Confidence interval
CMR: Cardiac magnetic resonance imaging
CVD: Cardiovascular disease
DLB: Dementia with Lewy bodies
HER: Electronic health record
FTD: Frontotemporal dementia
AnyDem: Any dementia
GWAS: Genome-wide association study
HR: Hazard ratio
ICD-10: International Classification of Diseases, 10th Revision
IVW: Inverse-variance weighted
LD: Linkage disequilibrium
LDL-C: Low-density lipoprotein cholesterol
MICE: Multiple imputation by chained equations
MR: Mendelian randomization
MR-Egger: Mendelian randomization Egger regression
NHGRI-EBI: National Human Genome Research Institute–European Bioinformatics Institute
NHS: National Health Service
OR: Odds ratio
OW: Overlap weighting
PS: Propensity score
SMD: Standardized mean difference
SNP: Single nucleotide polymorphism
UKB: UK Biobank
HMG-CoA: reductase 3-hydroxy-3-methylglutaryl-coenzyme A reductase
NPC1L1: Niemann-Pick C1-Like 1

## Data Availability Statement

The computer source code for this study is available at https://github.com/MIILab-MTU/HeartBrain_LDL_DementiaRisk.

## Acknowledgements

This research has been conducted using the dataset from NIH All of Us Research Program. We gratefully acknowledge the All of Us participants for their contributions, without whom this research would not have been possible.

